# DNA Methylation Analysis Identifies Clinically Relevant Lung Adenocarcinoma Subgroups

**DOI:** 10.1101/2024.08.26.24312568

**Authors:** Oluwadamilare I. Afolabi

**Affiliations:** ARL Bio, Ltd., Lagos, Nigeria

**Keywords:** Lung cancer, Cancer genomics, Biomarkers, Methylation

## Abstract

Lung adenocarcinoma (LUAD) is the most prevalent subtype of lung cancer and is characterized by significant molecular heterogeneity and poor prognosis, primarily due to late-stage diagnoses. Therefore, detailed molecular characterization of LUAD is crucial for developing biomarkers to accurately detect the disease in its early stages. This study investigates the role of DNA methylation in LUAD, emphasizing its potential as a biomarker for cancer detection and as a tool for understanding tumor biology. The study identified 4,925 differentially methylated sites (DMSs) and prioritized the top 200 DMSs for downstream analyses. Functional enrichment analysis revealed that site-specific hypermethylation in exon 1 and distal promoter regions are linked to critical developmental processes, including morphogenesis, pattern specification, stem cell differentiation, and synaptic transmission, suggesting that these epigenetic changes may disrupt normal cellular functions and contribute to tumorigenesis. Support vector machines demonstrated the diagnostic potential of these hypermethylated sites, achieving perfect classification of LUAD and normal adjacent tissues with as few as five features. Additionally, the strong correlation between methylation levels and feature importance scores further explained the predictive accuracy of these methylation markers. The study also identified distinct methylation subgroups within LUAD tumors, independent of traditional staging, each associated with unique transcriptional dysregulation and biological processes, such as DNA repair, immune response, and ribosome biogenesis. These findings not only enhance our understanding of LUAD pathophysiology but also underscore the clinical utility of DNA methylation as a diagnostic tool and guide for patient management.

## Introduction

Lung cancer remains the most prevalent and lethal malignancy, with non-small cell lung cancer (NSCLC) constituting approximately 85% of lung cancer cases [1, 2]. Despite advances in therapeutic strategies, the five-year survival rate for lung cancer remains low, underscoring the need for improved diagnostic and therapeutic approaches. Among NSCLC subtypes, lung adenocarcinoma (LUAD) is the most prevalent and is particularly challenging to treat due to its marked heterogeneity [3]. This heterogeneity represents a broad spectrum of molecular and histopathological variations, contributing to differences in tumor behavior and responses to therapy [4, 5]. Therefore, detailed molecular characterizations of LUAD are crucial for improved diagnosis and prediction of disease progression, enabling the development of more effective, tailored treatment strategies.

DNA methylation, specifically the addition of a methyl moiety to the cytosine residues of cytosine-guanine (CpG) dinucleotides, has emerged as a critical player in cancer biology. Regulatory elements within exons, untranslated regions (UTRs), promoters, and other genomic regions are important methylation sites, directly controlling gene expression at the transcriptional level [6, 7]. Aberrant methylation of these regions often results in the silencing of tumor suppressors or the activation of oncogenes, contributing to tumorigenesis and cancer progression. These epigenetic changes have been implicated in tumor complexity, aggressiveness, and therapeutic potential [8, 9, 10]. Thus, DNA methylation not only serves as an indicator of disease state but also represents an actionable target for therapy, making it invaluable for diagnostic and therapeutic applications.

This study leveraged array-based methylation data from The Cancer Genome Atlas (TCGA) to explore the DNA methylation landscape of LUAD. The study assessed the potential of the methylation statuses of exon 1 and distal promoter regions as biomarkers for disease detection and classification. The study findings highlight distinct methylation signatures correlating with LUAD subgroups, reflecting underlying biological differences that could be harnessed for personalized therapies. This work not only furthers our understanding of LUAD pathophysiology but also opens new avenues for developing methylation-based diagnostic tools, offering hope for improved management of this challenging disease.

## Materials and Methods

### Data acquisition and preprocessing

DNA methylation array data for 175 subjects diagnosed with LUAD (dbGaP accession phs000178) were retrieved from the TCGA database [11]. The dataset comprised genome-wide methylation profiles generated using the lllumina HumanMethylation450 BeadChip array, covering over 450,000 CpG sites across the human genome [12]. Matched normal adjacent tissue samples were also obtained for 31 of these subjects. The clinical characteristics and risk exposures of subjects in the TCGA LUAD cohort are summarized in Table 1. The absence of confounding factors was confirmed using T-tests for continuous variables and Chi-squared tests for discrete variables (Table 1).

**Table 1.** Clinical features of subjects in the TCGA LUAD cohort.

| Group | Tumor (N = 175) | Normal Adjacent (N = 31) | Statistic | p-value |
| --- | --- | --- | --- | --- |
| Age |  |  |  |  |
| 33.34 - 87.11 | 64.18 ± 10.3 | 64.53 ± 10.05 | t = 0.179 | 0.858 |
| Sex |  |  |  |  |
| Female | 105 (60%) | 19 (61%) | $\chi^2 = 0$ | 1 |
| Male | 70 (40%) | 12 (39%) |  |  |
| Stage |  |  |  |  |
| Stage I | 102 (58%) | 17 (55%) | $\chi^2 = 1.889$ | 0.756 |
| Stage II | 39 (22%) | 8 (26%) |  |  |
| Stage III | 21 (12%) | 2 (6%) |  |  |
| Stage IV | 10 (6%) | 3 (10%) |  |  |
| Unreported | 3 (2%) | 1 (3%) |  |  |
| Cigarettes per Day |  |  |  |  |
| 0 - 8.44 | 1.72 ± 1.69 | 1.61 ± 1.67 | t = -0.328 | 0.745 |

### Differential methylation analysis

To identify CpG sites differentially methylated in LUAD, a subset comprising one-third of the LUAD cohort was randomly selected (Table S1) and analyzed using DESeq2 v1.42.1 [13]. DESeq2, widely used for the analysis of count-based data to identify differentially expressed genes, was adapted in this study to identify differentially methylated sites (DMSs). Before differential methylation analysis, CpG sites with consistently low methylation levels, specifically those withiS’-values less than 0.1 in all samples of the smallest sample group, were filtered out to minimize noise. The 200 most differentially methylated sites, based on absolute fold change and a false discovery rate (FDR) adjusted p-value threshold of 0.01, were prioritized for further analyses. The overall methylation changes of these DMSs were evaluated using a volcano plot, which depicts the relationship between fold change and statistical significance. Generally, volcano plots provide a clear visualization of the extent and direction of methylation differences. Additionally, the distribution of DMSs across various genomic regions was compared using a bar plot to assess which genomic regions were more prone to differential methylation in LUAD. Furthermore, samples were clustered by methylation patterns using hierarchical clustering with Ward’s method and visualized using heatmaps [14]. Ward’s method minimizes the total within-cluster variance, ensuring that the resulting clusters are as homogeneous as possible [15], allowing for a clear representation of the relationships between samples based on their methylation profiles.

### Functional enrichment analyses

DMSs were first linked to their associated gene. The OMS-associated genes were then stratified by the genomic location of their DMSs and subjected to Gene Ontology (GO) enrichment analysis using the clusterProfiler v4.10.1 [16]. This analysis aimed to identify biological processes significantly overrepresented among the genes associated with DMSs in specific genomic regions. A q-value threshold of 0.05 was applied to minimize the likelihood of false positives. GO enrichment analysis was performed on genes uniquely dysregulated in methylation clusters identified during the study to further explore the functional implications of methylation patterns. This analysis provided insights into the distinct biological processes that may be driving the behavior of each methylation-defined LUAD subgroup. The top 10 enriched GO processes were visualized using dot plots for all functional enrichment analyses.

### Machine learning

LUAD cohort samples were split into training (60%) and test (40%) sets to build and evaluate predictive models. Support vector machine (SVM) models were trained on the training set using various feature sets, specifically the most methylated 1, 5, 10, or all exon 1 or distal promoter DMSs [17]. Several key metrics were extracted using caret v6.0-94 [18] and pROC vl.18.5 [19] to evaluate the performance of these models on the test set. Detailed breakdowns of true positives, true negatives, false positives, and false negatives were visualized in a confusion matrix. ROC curves were further generated for each model to assess their classification performance. Area Under the Curve (AUC) values were reported with 95% confidence intervals (Cl) as a summary measure of model accuracy, providing a robust metric for evaluating the performance and reliability of the SVM models. Using a random forest algorithm [20], importance scores, quantifying each DMS’s contribution to reducing prediction uncertainty across all decision trees, were calculated. These importance scores were then correlated with average methylation levels of the corresponding DMSs to explore potential relationships. Correlation was assessed by fitting a linear regression model, with Pearson’s correlation coefficients (R) and p-values reported.

### Gene expression analysis

To detect gene dysregulation within LUAD methylation subgroups, gene expression quantification data for four subjects per methylation cluster (dbGaP accession phs000178) were retrieved from the TCGA database [11]. Genes with less than ten counts in all samples of the smallest sample group were filtered out to reduce noise. Differentially expressed genes (DEGs) were then identified between the methylation clusters 2, 3, 4, and 5 and cluster 1 (normal adjacent tissue) samples at an FDR-adjusted p-value threshold of less than 0.05. To further emphasize the uniqueness of these clusters, overlaps in the sets of upregulated and downregulated genes among clusters 2, 3, 4, and 5 were visualized using upset plots [21]. This visualization approach allowed for a clear representation of shared and unique dysregulated genes across the clusters, highlighting both common pathways and cluster-specific gene expression changes that may contribute to the distinct biological behaviors of each subgroup.

## Results

### Aberrant hypermethylation in LUAD

The differential methylation analysis revealed that 2,875 CpG sites (24%) were hypermethylated, while 2,050 sites (17%) were hypomethylated in LUAD tumors compared to normal adjacent tissue samples (Table S2). For downstream analyses, the top 200 differentially methylated sites (DMSs) by fold change and FDR-adjusted p-values were selected, all of which exhibited above four-fold hypermethylation (Fig. 1A). Notably, the majority of these top DMSs were located in exon 1 (55 sites), closely followed by the distal promoter (TSS1500) region (46 sites). In contrast, the 3’ UTR (2 sites) had the fewest top DMSs (Fig. 1B).

**Fig. 1.**
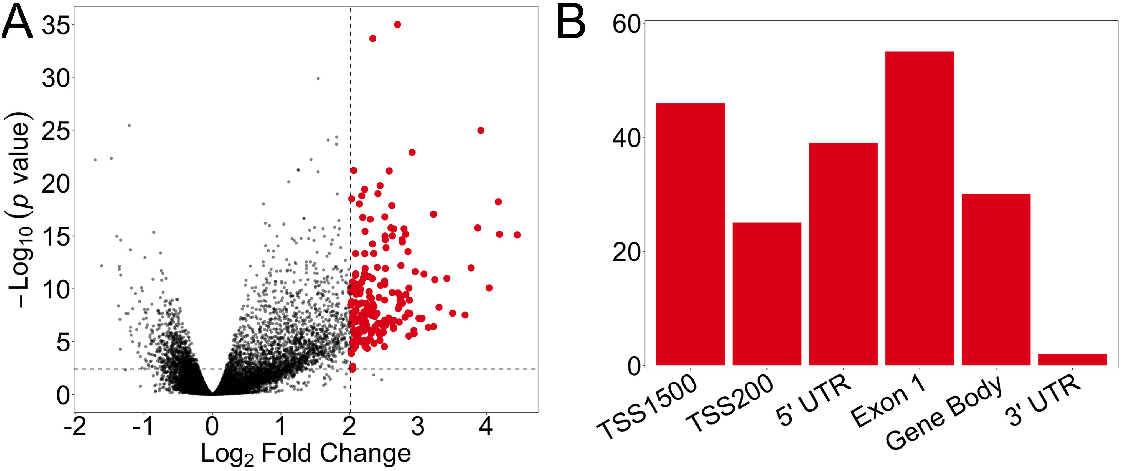
Differential methylation in LUAD. **(A)** Volcano plot depicting DMSs in LUAD versus NAT samples. The top 200 DMSs, based on absolute fold change and an FDR-adjusted p-value threshold of 0.01, are highlighted in red. **(B)** Bar plot illustrating the distribution of hypermethylated CpG sites across various genomic regions.

Functional enrichment analysis of DMSs in LUAD revealed several biological processes significantly associated with hypermethylation in different genomic regions (Table S3). The analysis highlighted that genes near hypermethylated CpG sites in exon 1 regions are involved in critical developmental processes such as pattern specification process, stem cell differentiation, and regulation of neuron differentiation (Fig. 2A). These processes suggest that the hypermethylation observed may contribute to the disruption of normal development and differentiation pathways in LUAD, potentially contributing to tumorigenesis. In 5’ UTRs, significant enrichment was also observed for processes related to morphogenesis (Fig. 2B). This could indicate a link between aberrant methylation in these regulatory regions and the altered expression of genes crucial for proper development, which might contribute to the abnormal growth in LUAD tumors. For DMSs in distal promoter regions, the enriched biological processes were similarly associated with the morphogenesis of branching structures, but also the regulation of synaptic potential and chemical synaptic transmission (Fig. 2C). The involvement of these processes underscores the potential role of epigenetic modifications in altering epithelial and neural signaling pathways, which could influence tumor behavior.

**Fig. 2.**
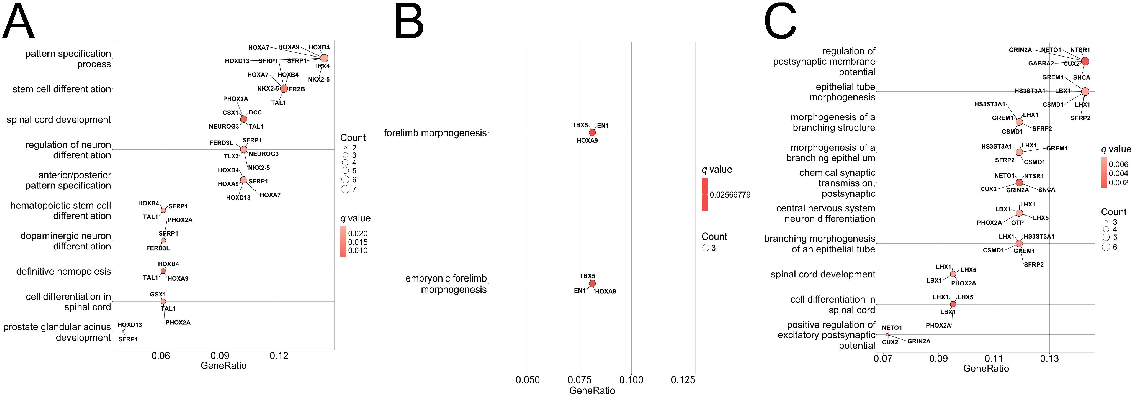
Biological processes associated with hypermethylation in LUAD. Dot plots highlighting biological processes associated with differential methylation of **(A)** exon 1, **(B)** 5’ UTR, and **(C)** distal promoter regions.

### Diagnostic potential of exon1 and distal promoter methylation

Since exon 1 and distal promoter region, hypermethylation appeared to be the most biologically relevant, these regions were prioritized for further analyses. Heatmaps of the methylation levels of these DMSs, with samples sorted vertically by average CpG site methylation and CpG sites sorted horizontally by average sample methylation, were generated. These heat maps confirmed that the selected CpG sites were hypermethylated exclusively in tumor samples, suggesting their potential as biomarkers for LUAD (Fig. 3A-B). Some CpG sites exhibited more consistent hypermethylation within tumor samples than others, indicating a complex yet patient-specific methylation pattern.

**Fig. 3.**
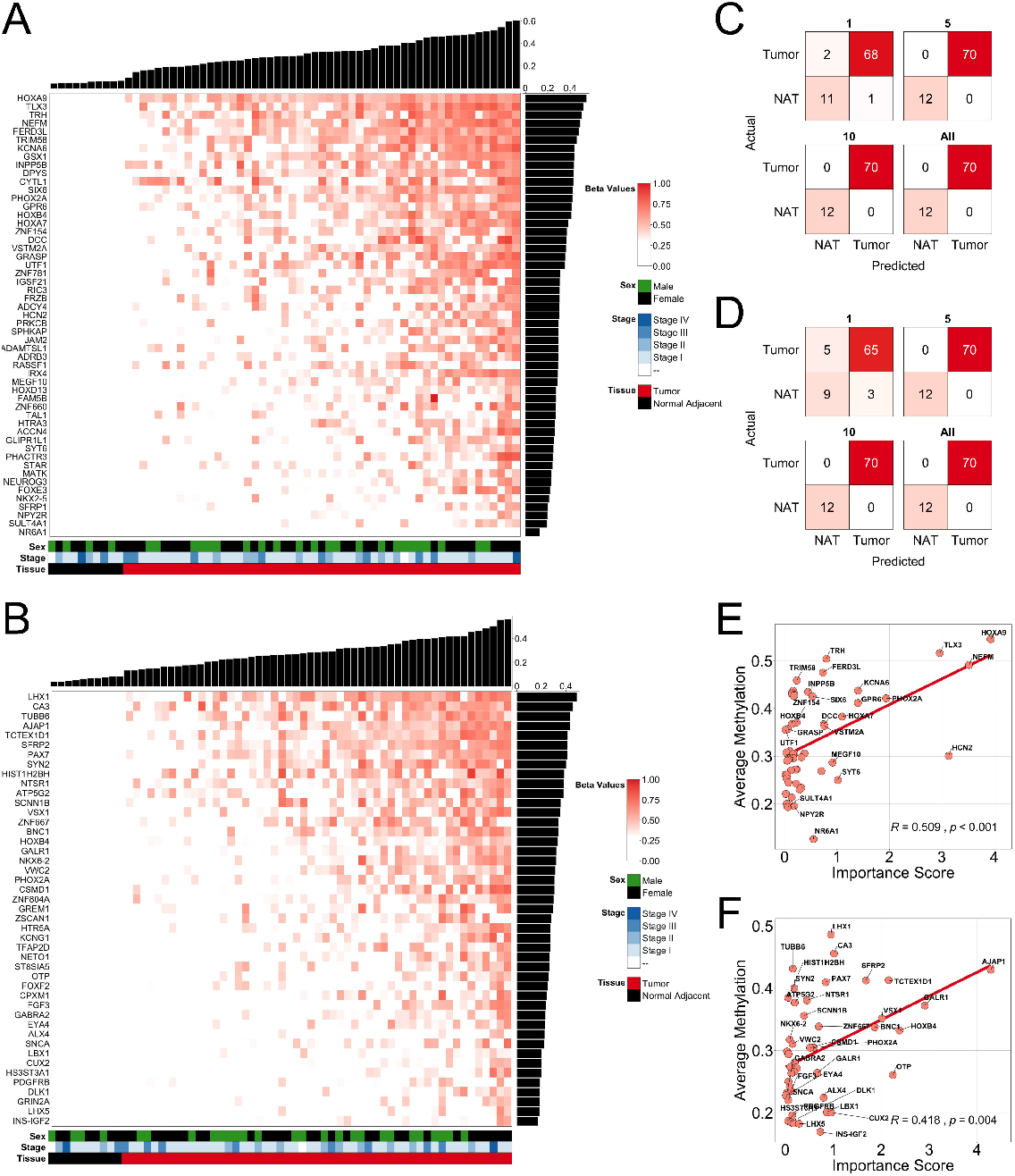
Exon 1 and distal promoter methylation as classifiers for LUAD. Heatmaps showing methylation patterns in NAT and LUAD samples for **(A)** exon 1 and **(B)** distal promoter regions. Vertical bar plots represent average methylation levels across samples, while horizontal bar plots show average CpG site methylation levels. Confusion matrices depicting the performance of SVM models trained and tested on 1, 5, 10, and all DMSs from **(C)** exon 1 and **(D)** distal promoter regions. Correlation plots of average methylation and feature importance for **(E)** exon 1 and **(F)** distal promoter DMSs in random forest models.

The efficacy of these DMSs as classifiers was validated using support vector machine (SVM) models, which were trained on varying numbers of DMSs from exon 1 (Fig. 3C) and distal promoter (Fig. 30) regions. Remarkably, just five DMSs were sufficient to achieve perfect classification of samples into tumor and normal adjacent tissue. A model trained on a single exon 1 OMS with the highest average methylation (cg26521404 in the HOXA9 gene) achieved an AUC of 0.99 (95% Cl, 0.97-1.00) (Fig. S1A). However, the model trained on the distal promoter OMS with the highest average methylation (cg22660578 in the LHX1 gene) yielded a slightly lower AUC of 0.94 (95% Cl, 0.89-0.99) (Fig. S1B). Furthermore, random forest feature importance scores correlated significantly with average methylation levels for both exon 1 (R = 0.509, p < 0.001) and distal promoter regions (R = 0.418, p < 0.004), underscoring the impact of consistent methylation at these sites on prediction accuracy (Fig. 3E-F).

### LUAD tumors cluster into distinct methylation subgroups

Unsupervised hierarchical clustering of tumor and normal adjacent tissue samples based on methylation patterns in exon 1 and distal promoter regions identified five distinct methylation subgroups (Fig. 4). Cluster 1 exclusively comprised of normal lung tissue samples, while Clusters 2-5 were comprised entirely of tumor tissue samples. Cluster 2 had the lowest proportion of male patients (20%), while Cluster 5 had the highest (68.75%). Cluster 3 exhibited the least smoking history among patients and showed the lowest levels of methylation, suggesting a potential link between smoking and overall methylation levels. There was no apparent clustering of samples based on cancer stages, indicating that methylation subgroups may be independent of traditional staging.

**Fig. 4.**
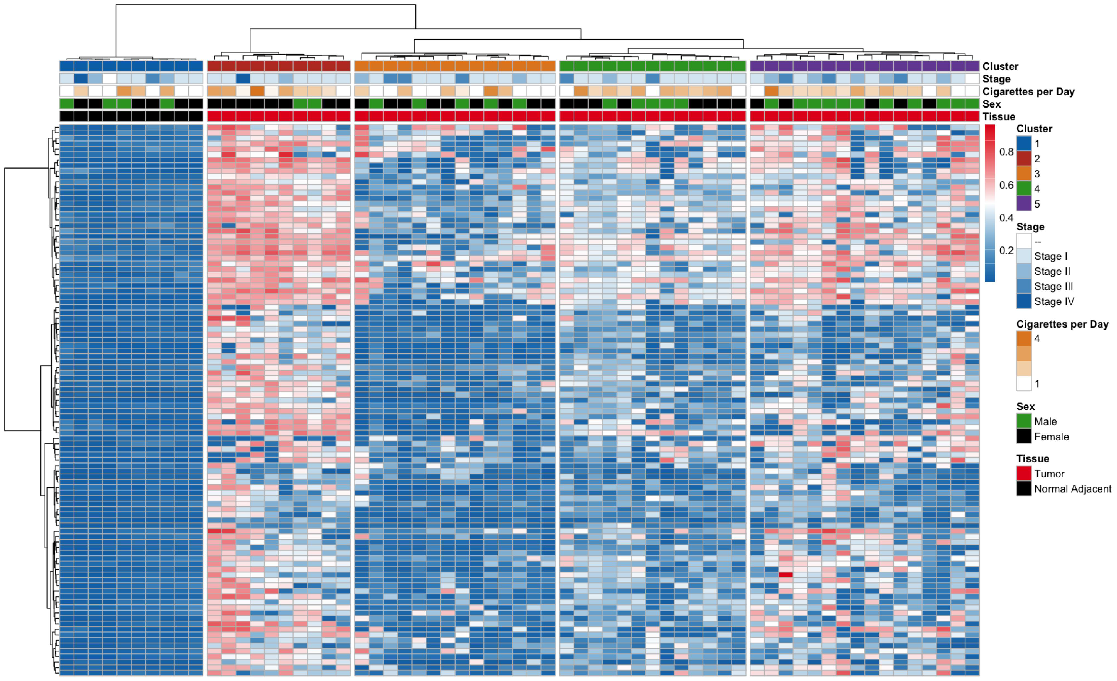
Methylation subgroups identified in LUAD. Heatmap of NAT and LUAD samples clustered by methylation patterns in exon 1 and distal promoter regions.

### Gene dysregulation within methylation subgroups

Gene expression analysis across the methylation-defined LUAD subgroups revealed significant dysregulation patterns specific to each cluster. Fig. SA and 5B illustrate the overlaps of genes that are either upregulated or downregulated across Clusters 2, 3, 4, and 5, compared to normal adjacent tissues. Cluster 3 exhibited a distinct profile with the highest number of uniquely upregulated genes, which may be attributed to its relatively lower methylation levels. Enrichment analyses further identified distinct biological processes associated with the differentially expressed genes within these clusters (Fig. 5C-F). For example, Cluster 3 was strongly associated with DNA repair processes, particularly those involved in homologous recombination and replication fork processing (Fig. 5D). In contrast, Cluster 4 was enriched for immune-related processes, such as leukocyte activation and chemotaxis (Fig. SE). Additionally, Cluster 5 displayed enrichment in processes related to ribosome biogenesis and translation, indicating high transcriptional activity (Fig. SF). Conversely, Cluster 2 was associated with only one biological process, suggesting these tumors are less distinct in their transcriptional profile compared to the other clusters (Fig. SC). These findings underscore the molecular heterogeneity of LUAD, with most methylation subgroups exhibiting distinct transcriptional programs, which could be leveraged for more tailored therapeutic strategies.

**Fig. 5.**
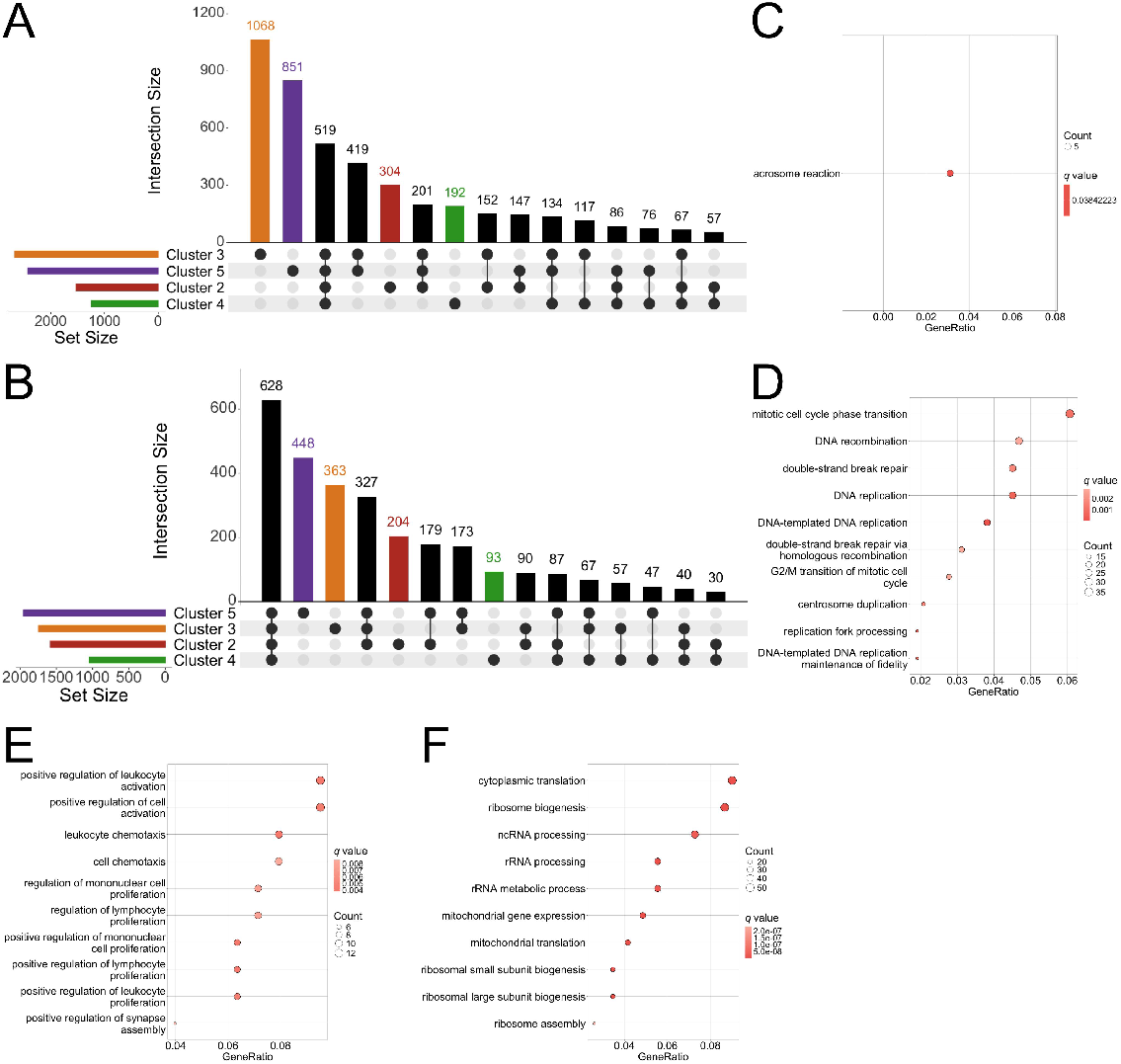
Biological processes associated with LUAD subgroups. Upset plots showing overlaps of **(A)** upregulated and **(B)** downregulated genes in clusters 2, 3, 4, and 5 versus NAT samples. **(C-F)** Dot plots highlighting biological processes associated with clusters 2, 3, 4, and 5, respectively.

## Discussion

Lung cancer remains the deadliest malignancy worldwide, primarily due to late-stage diagnoses [1, 22]. The challenge of early detection is exacerbated by the heterogeneous nature of lung cancer, which often leads to diagnostic delays and suboptimal treatment outcomes [23]. Despite therapeutic advances, the prognosis for lung cancer patients remains poor, underscoring the need for biomarkers capable of accurately detecting the disease in its early stages. Several studies have highlighted DNA methylation’s promise as a cancer detection biomarker [24, 25, 26]. This study’s findings emphasize the critical role of DNA methylation in LUAD and demonstrate the potential of high-resolution DNA methylation profiling for accurate cancer detection and characterization. This study identified many CpG sites with aberrant hypermethylation in LUAD tumors, particularly in exon 1 and distal promoter regions. The overrepresentation of hypermethylated CpG sites in these regions suggests they may play a crucial role in modulating gene expression, thereby contributing to LUAD pathogenesis.

Functional enrichment analysis of genes associated with these CpG sites further elucidated the biological implications of their hypermethylation. The association of exon 1 hypermethylation with developmental processes such as pattern specification, stem cell differentiation, and neuron differentiation highlights a potential mechanism by which epigenetic alterations disrupt normal cellular differentiation and contribute to tumorigenesis. These findings are consistent with previous research linking exon 1 and promoter hypermethylation to the silencing of tumor suppressor genes and activating oncogenic pathways [27, 28, 29]. Similarly, the hypermethylation observed in 5’ UTRs and distal promoter regions, linked to processes like morphogenesis and synaptic transmission, suggests a broader impact of DNA methylation on epithelial and neural signaling pathways in LUAD. These findings align with the growing recognition of epigenetic modifications as key cancer progression drivers and potential therapeutic targets [30, 10].

The diagnostic potential of exon 1 and distal promoter hypermethylation was further validated by SVM models, which achieved perfect classification of LUAD and normal adjacent tissues using as few as five features. Moreover, the strong correlation between feature importance scores and average CpG site methylation levels underscores the link between the accurate LUAD prediction observed and the consistent methylation of these sites in tumor samples. The use of machine learning models in methylation analysis has been increasingly recognized for its ability to enhance the predictive power of epigenetic biomarkers in cancer diagnostics [31]. Additionally, the identification of distinct methylation subgroups within LUAD tumors, independent of traditional staging, highlights the molecular heterogeneity of this cancer type and suggests that methylation profiling could provide a more nuanced understanding of tumor biology and ultimately inform personalized treatment strategies.

Finally, the gene expression analysis across the methylation-defined subgroups revealed significant transcriptional dysregulation, with each cluster associating with unique biological processes. These findings align with previous studies that have demonstrated how distinct methylation patterns correlate with differential gene expression, influencing tumor behavior and treatment response [32, 33]. The distinct profiles observed, particularly the enrichment of DNA repair processes in Cluster 3, immune-related processes in Cluster 4, and ribosome biogenesis in Cluster 5, provide compelling evidence of the potential for these methylation subgroups to inform targeted therapeutic approaches. Overall, this study deepens our understanding of LUAD pathophysiology and underscores the clinical utility of DNA methylation as both a diagnostic tool and a guide for personalized patient management.

## Data Availability

All the data used in this study are openly available in the TCGA Genomic Data Commons, dbGaP accession phs000178.

https://portal.gdc.cancer.gov

## Acknowledgments

The results shown here are based on openly available human data from the TCGA Research Network: https://www.cancer.gov/ccg/research/genome-sequencing/tcga.

## Ethics Approval and Consent

Not applicable.

## Data Availability

All the data used in this study are openly available in the TCGA Genomic Data Commons at
https://portal.gdc.cancer.gov, dbGaP accession phs000178.

