## Supplemental Fig. S1 for "DNA Methylation Analysis Identifies Clinically Relevant Lung Adenocarcinoma Subgroups"

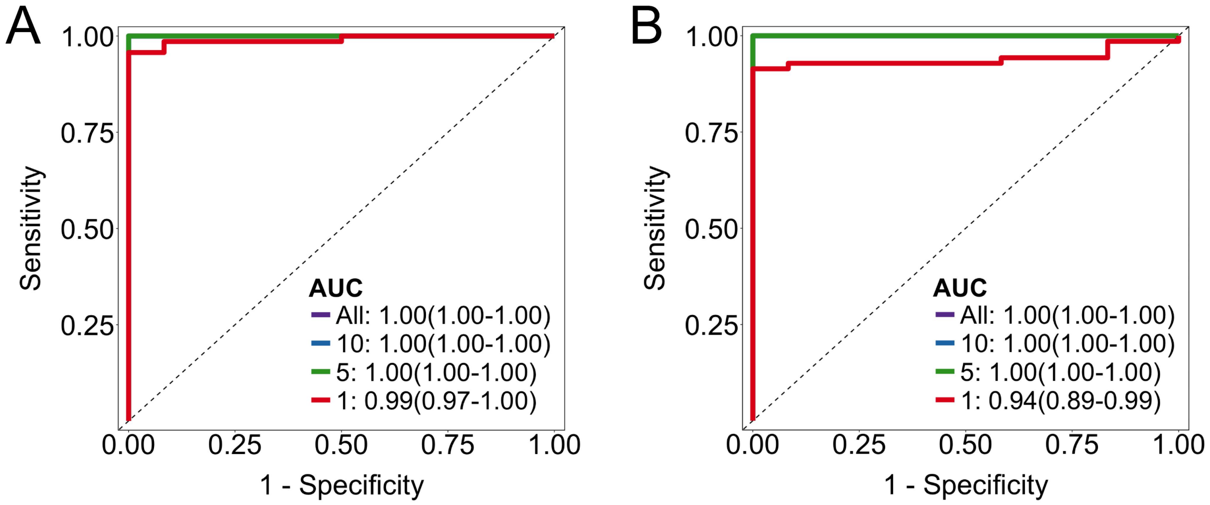


**Fig. S1. Exon 1 and distal promoter methylation as classifiers for LUAD.** ROC curves illustrating the performance of SVM models trained on 1, 5, 10, and all (**A**) exon 1 and (**B**) distal promoter DMSs.
